# Analyzing Surgical Volumes, Rates, and Need in Rural India

**DOI:** 10.1101/2021.11.03.21265903

**Authors:** Siddhesh Zadey, Joao Ricardo Nickenig Vissoci

## Abstract

**Background:** Globally, 5 billion people lack timely access to safe and affordable surgical care, with over a fifth of them living in India. Solving India’s surgical access issues can have high returns on investment. While healthcare access and unaffordability problems are well-known in India particularly among its rural people, research on surgical volumes and need is scant. This study attempts to fill the research gap through high-resolution nationwide estimates that have direct implications for India’s national surgical plan.

**Methods:** Secondary data analysis with a diverse geospatial and statistical toolbox was used to create the national, state, and district-level estimates for surgical rates and c-section proportions and their corresponding met need w.r.t. to the globally prescribed thresholds – 5000 major surgeries (those requiring anesthesia) per 100,000 (Lancet Commission on Global Surgery) and 10-15% of all institutional deliveries (World Health Organization).

**Results:** Nationally, only 6.81% of need for major surgical operations was met for rural India. 13.6% of the institutional deliveries were c-sections falling within the WHO-prescribed range of 10-15%. There were marked variations at state and district-levels and significant rural-urban differences for surgical rates and c-section proportions. We validate our estimates based on data from Health Management and Information System against existing sources that are commonly used in academic and policy research.

**Conclusions:** Our methodological workflow has high translational value for global surgery research in low-and-middle-income countries. For India, these are the first such nationwide findings that can direct the development of a National Surgical, Obstetric, and Anesthesia Plan (NSOAP).

## 1. Introduction

In 2015, the Lancet Commission on Global Surgery (LCoGS) reported that 5.3 billion people globally lack timely access to safe and affordable surgical care (1, 2). Of these, 3.3 billion people with no access to surgical care reside in low-and-middle-income countries (LMICs). In relative terms, 99.3% and 96.7% of people in low- and lower-middle-income countries lack access compared to 68.3% and 26.4% in upper-middle- and high-income countries. The disparity in access to essential and emergency surgery (3) is thought to be responsible for 4.7 million avertable deaths in LMICs (4). The Commission proposed six global surgery indicators that included the surgical volumes per 100,000 people among others (5). The proposed implementation solution to the problem was was the proposal of National Surgical, Obstetric and Anesthesia Plans (NSOAPs) that could bring political attention to surgical care issues and advocate for the incorporation of surgery in the national health policy agenda (6). Recently, NSOAPs have been proposed for Zambia, Rwanda, and others to direct evidence-based participatory national surgical system strengthening (7). A quintessential step in developing an NSOAP is a comprehensive country-wide assessment of the surgical care system to benchmark the baseline and determine target achievement (7, 8).

### 1.1 Evidence on Surgical Volumes, Rates, and Need in India

#### 1.3.1 Insights from the LCoGS

Research accompanying LCoGS generated country-level data for access dimensions among several other variables as summarized in **Table 1**. In some instances, the data were aggregated at a higher level corresponding to World Bank Income Groups or Global Burden of Diseases (GBD) Regions. Erring on the higher side, only one-fifth of surgical need was met among the Indian population. India also depicted deficits in surgical volumes/rates and workforce. Further, it was estimated that at the current levels, it would be well beyond 2030 to reach the targeted surgical rates, denoting the need to integrate surgical care in the SDGs-2030. When compared to forgone macroeconomic losses, it is evident that investments in scaling up surgical care are beneficial.

**Table 1:**
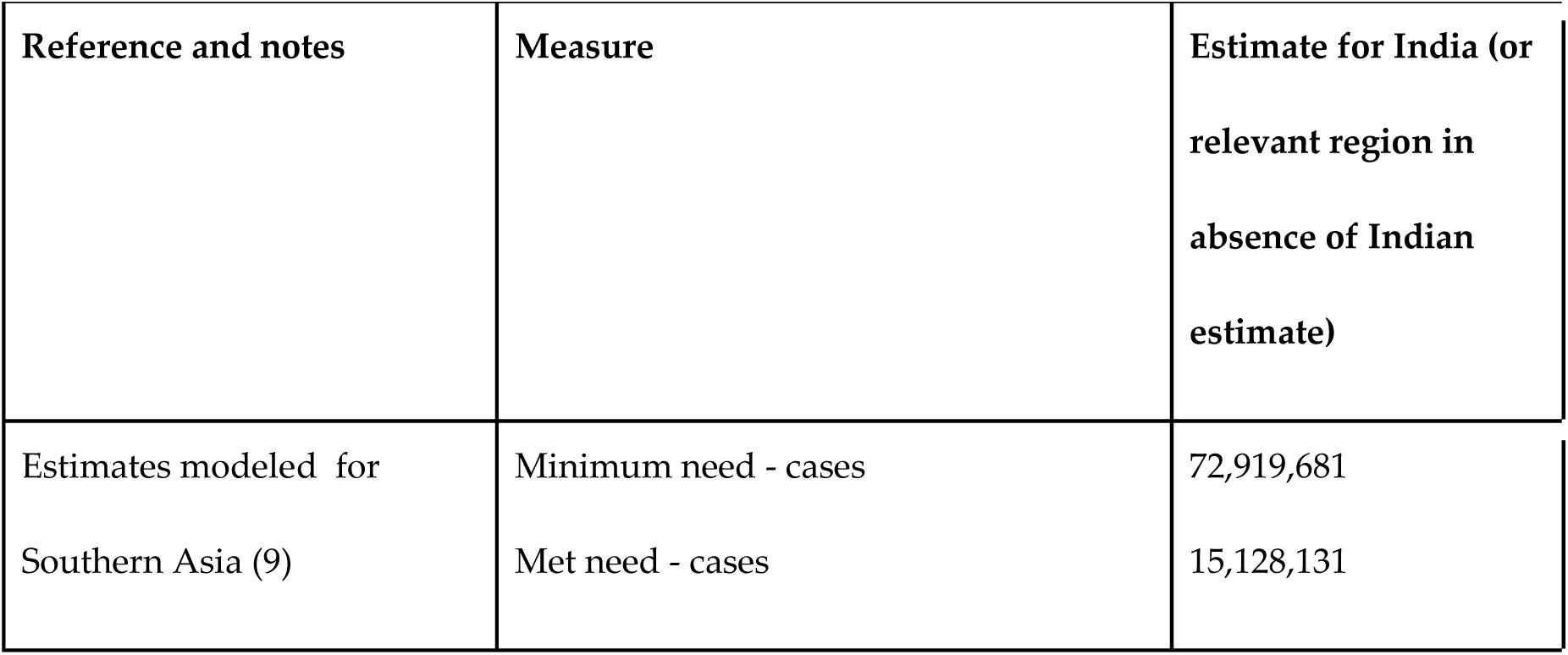
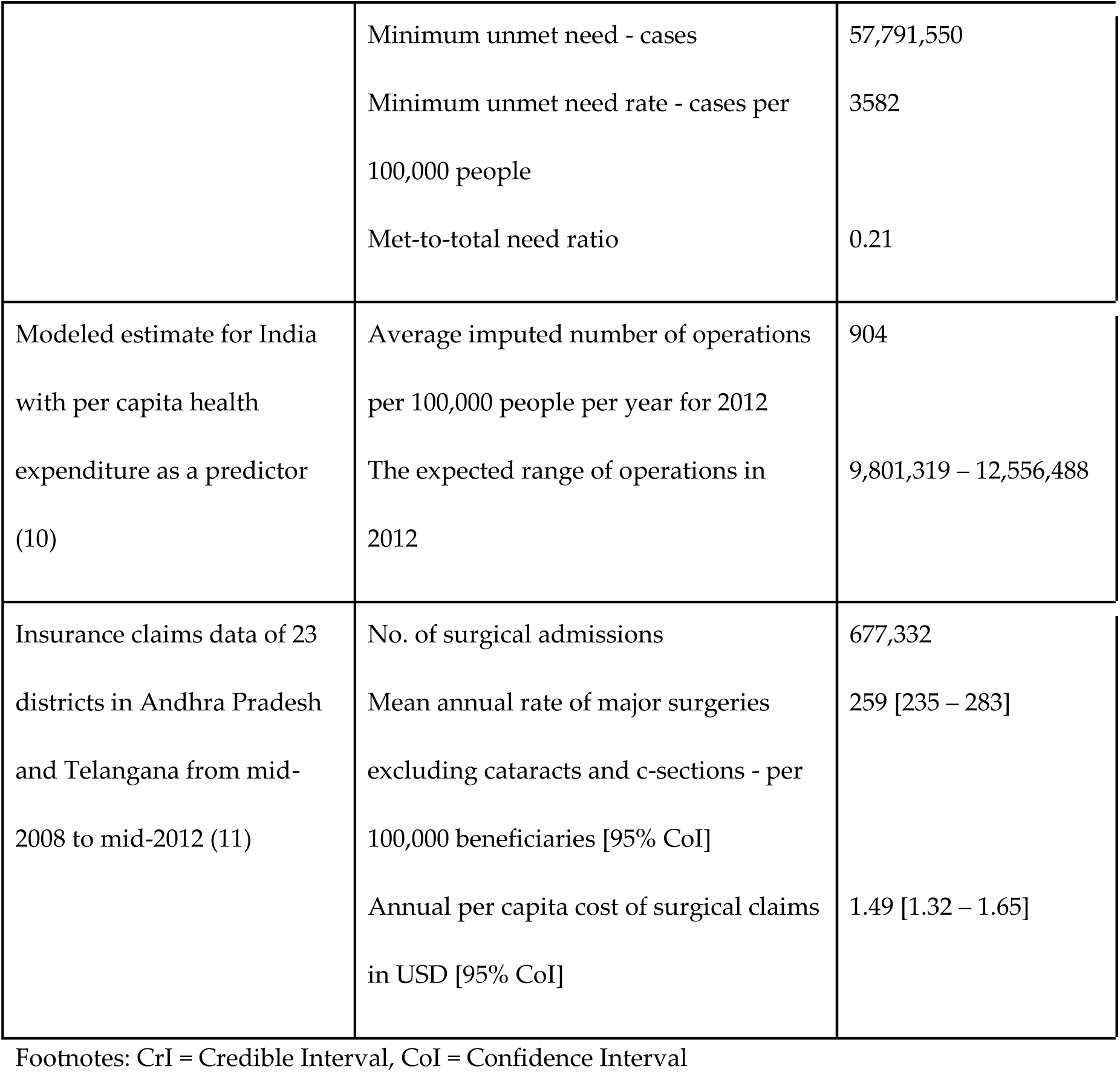
Surgical Capacity Dimension and Surgical Care Indicator 3 estimates reflecting surgical volumes, rates, and need relevant to India from LCoGS (2015)

| Reference and notes | Measure | Estimate for India (or relevant region in absence of Indian estimate) |
| --- | --- | --- |
| Estimates modeled for Southern Asia (9) | Minimum need - cases | 72,919,681 |
|  | Met need - cases | 15,128,131 |
|  | Minimum unmet need - cases | 57,791,550 |
|  | Minimum unmet need rate - cases per 100,000 people | 3582 |
|  | Met-to-total need ratio | 0.21 |
| Modeled estimate for India with per capita health expenditure as a predictor (10) | Average imputed number of operations per 100,000 people per year for 2012 | 904 |
|  | The expected range of operations in 2012 | 9,801,319 – 12,556,488 |
| Insurance claims data of 23 districts in Andhra Pradesh and Telangana from mid-2008 to mid-2012 (11) | No. of surgical admissions | 677,332 |
|  | Mean annual rate of major surgeries excluding cataracts and c-sections - per 100,000 beneficiaries [95% CoI] | 259 [235 – 283] |
|  | Annual per capita cost of surgical claims in USD [95% CoI] | 1.49 [1.32 – 1.65] |

Footnotes: CrI = Credible Interval, CoI = Confidence Interval

The LCoGS estimates were novel and fill in important gaps in the global surgery research, particularly for LMICs. Even so, in the Indian context, these estimates have limitations. Most of the data used for the estimates dated back to the late 2000s or early 2010s (2012 - for surgical volumes (10)). Further, data was not collected but imputed through modeled approximations for India (9). No subnational estimates were presented. Post-LCoGS, the Global Surgical Care Indicators Initiative reports (12, 13) regularly update the six indicators. However, the two reports (2015 and 2017) published yet contain no new data on surgical volumes or need in India.

Regardless of the limitations at the research end, the Commission generated enough momentum to bring together several national actors in India that could push forward the surgical care agenda. The Karad Consensus Statement (KCS) (2015) (14) framed initially by the Association of Rural Surgeons of India (ARSI) (http://www.arsi-india.org/) was endorsed by several stakeholders at the National Indian Surgical Forum (2016) (15). KCS identified that strengthening rural surgical systems, expanding and optimizing the surgical workforce, and tackling the blood deficit, particularly for rural locations should be a high priority for India. Subsequently, the ‘Implementing The Lancet Commission on Global Surgery in India’ (i-LCoGS-India) provided the action items towards sustainable resolution of the above-mentioned problems (16). Backed by the Commission and WHO and driven by local stakeholders from diverse backgrounds, i-LCoGS-India has two Secretariats in New Delhi and Mumbai and a field office in Bihar to facilitate the administrative and research goals arising from the ARSI-LCoGS-others collaboration. Access to surgical care in rural India is unequivocally the highest policy priority and in turn, requires urgent research focus.

#### 1.3.2 Other evidence

Apart from the LCoGS, the Disease Control Priorities Network (DCPN) in its 3^rd^ volume (DCP3) (2015) focused on essential surgery (3). DCP3 defined essential surgical conditions as that – a) are primarily or extensively treated by surgical procedures and other surgical care, b) have large health burden, and c) can be successfully treated by a surgical procedure and other surgical care that is cost-effective and feasible to promote globally. Surgical programs and packages catering to essential surgical conditions formed essential surgery. DCP3 provided comprehensive evidence on the cost-effectiveness and benefits of surgical scale-up in LMICs. Particularly for India, it reviewed the literature for various surgical platforms including cataracts, mosquito-net mesh hernia repair, etc., demonstrated that high benefit-to-cost ratio (BCR) of cleft-lip surgical repairs, and pointed to access problems in rural places due to surgical setup unavailability at health facilities lower than the district hospitals in the referral hierarchy.

Apart from the LCoGS and DCP3, small-to-medium scale regional studies have looked at surgical need in rural and urban areas (17–19). The national-level surgical need benchmark using data from a universal healthcare coverage (UHC) cohort (n = 88,273, Contributory Health Service Scheme (CHSS) members) was estimated to be 3646 procedures per 100,000 people (17). However, this cohort did not demographically and socioeconomically match the Indian population, excluded certain kinds of surgeries, and assumed no accessibility or acceptability issues. Two studies using the Surgeons OverSeas Assessment of Surgical Need (SOSAS) self-reports in low-income households in Ahmedabad city (n= 10,330 from 2066 households) (18) and rural Haryana (n= 93 from 50 households) (19) estimated the surgical need at 3.46% and 30.1% with unmet proportions at 42.33% and 6.5%, respectively. While useful, these estimates lack comprehensiveness for national-level planning and limited use for rural India. Hence, nationwide surgical access dimension estimates for India and specifically rural India are urgently needed to create a strong foundation for surgical planning.

### 1.2 The two ‘India-s’: Health in Rural Bharat

Regardless of growing urbanization, the most recent 2011 census revealed that about 833 million (68.84%) of the 1.21 billion Indians lived in rural areas (20). The colloquial saying is that India hosts two countries – the minority and materially-rich urban India and the majority and naturally-gifted rural *Bharat* (Hindi name for India). While India is steadily moving towards UHC, *Bharat* is thought to grapple for basic healthcare. The National Burden Estimates (NBE) demonstrated that, in 2017, rural India accounted for over 75% of all deaths and DALYs. The DALY rate (per 100,000) for all ages was 40,400 in rural areas compared to 27,400 in the urban counterparts (21). Among the top 15 causes of DALYs, rural areas have a greater burden of all causes except ischemic heart disease and musculoskeletal disorders, relative to urban areas. The stark rural-urban health differences can be attributed to a plethora of healthcare and socioeconomic differences and disparities.

Rural India faces deficits in access to healthcare (22), health infrastructure, and manpower (23) Further, the quality of health service delivery remains questionable in large parts of the rural public health system due to a lack of underlying resources and quality control processes. Hence, people in these areas often have to travel large distances to seek good-quality care in urban private hospitals forcing them to spend a larger proportion of their limited incomes on healthcare compared to the urban counterpart (24). As a result, rural households are at a significantly higher risk for CHE (25). Complementing the supply-side factors, the demand for health-seeking for rural households is also limited by lower education rates, higher unemployment, and greater poverty (26) compared to their urban counterparts, forcing the ‘rural poor’ to take the brunt (27). The problems of the rural health systems can only be expected to exacerbate for the rural surgical systems.

The National Rural Health Mission (NRHM) launched in 2005 (now subsumed under National Health Mission - NHM) has been instrumental in improving the rural public health infrastructure (28) and manpower (29), which, in turn, has enhanced accessibility and had some positive impact on affordability (30). To extend NRHM’s promise of UHC for rural India, the Ayushman Bharat (AB) program was launched in 2018. AB has two objectives targeted by one component each – to expand the primary healthcare system through the creation and scale-up of Health and Wellness Centers (AB-HWCs) and to initiate comprehensive social health insurance (SHI) under the Pradhan Mantri Jan Arogya Yojana (AB-PMJAY) for reducing OOP expenditure (OOPE) among the members of the deprived classes seeking good quality care at secondary and tertiary public and private hospitals (31). While it is too early to understand AB’s impact, NHM has contributed to significant improvements in the population-level health outcomes particularly for maternal and child health and communicable diseases (32). However, its contribution specifically to rural surgical care remains elusive, or largely unassessed.

It could be speculated that a large proportion of the rural DALYs could be surgically averted making ‘met surgical need’ an important target indicator of research and policy. Paradoxically, rural India is a neglected population in global surgery research although being of high interest in the Indian health policy and planning domains pointing to a chasm that needs urgent addressing.

### 1.3 Rural surgical care in India: Behind the hour

To ensure UHC for India, it is essential to bring UHC to its rural people. UHC cannot be achieved without adequately attending to surgical care, a critical and integral component of any healthcare system. A comprehensive NSOAP could lay out the roadmap and targets for rural surgery in India, however, such a plan would require robust nationwide evidence. As pointed before, there is negligible data on rural surgical care with no high-resolution nationwide assessment. India is running behind the hour when it comes to generating evidence to strengthen its rural surgical care. This study is directed towards filling the critical evidence gap. Our primary aim is to synthesize national and subnational (district and state-level) estimates for surgical rates, c-section proportions and their corresponding met need for rural populations of India for 2017-2018. Our secondary aim is to compare rural and urban counterparts to point to any existing disparities.

## 2. Methods

### 2.1 Data Sources

A significant study product of the current study is assimilating data from eclectic international, national, and subnational sources. Identification and compilation of relevant data sources and the methodological workflow for harmonization have high value for surgical care and health systems research in LMICs. **Table 2** presents a detailed list of data sources and data use considerations. Our secondary data assimilation workflow can be described as:

1. We reviewed data contingencies of high-level national health reports, census data, household surveys, nationally representative sample surveys, health registries, and the country’s health management information system (HMIS) to identify the required variables. While tedious, this step was critical to ensure that ‘most appropriate’ data are used to create the estimates. For instance, we decided to use HMIS over sample survey data for surgical volumes as HMIS is supposed to have wider coverage, regular (monthly) upkeep, and greater utility towards local health planners. Cross-dataset variable enlisting also points to variable overlaps and data collection heterogeneities. Similar variables recorded across different datasets can aid validation. For instance, we compared the c-section volumes across HMIS and the corresponding National Family Health Survey-4 (NFHS-4) estimates from another study (33), helping us validate HMIS data.
2. We assessed the data quality by reviewing literature previously citing the data sources. Understanding the source limitations early on helps in designing analyses that could accommodate limitations.
3. We assessed the highest possible geographic resolution and a common period across data sources for reliable large-scale situational analysis. Here, we identified districts to be geographic-administrative units of the highest possible resolution and the years 2017-2018 suitable for situational analysis. The HMIS data used for calculating surgical rates and c-section proportions was obtained for April 2017 to March 2018. The population estimates for 2017 were used for all calculations.

**Table 2:** Data sources used in the current analysis for India.

| Data source | Source information | Extracted data and considerations |
| --- | --- | --- |
| WorldPop (34)<br>(India files) | It is a global high-resolution geospatial population datasets library with well-characterized and validated estimates (35). | Raster (.tif) for UN-adjusted unconstrained population counts (people per pixel) at 1 km<sup>2</sup> resolution for India for 2017 (36). These were used for population estimation. |
| GADM version 3.6 (37) | It is a data repository for administrative boundaries | Shape boundary files (with associated polygon data frames) for |
| (Indian boundaries) | standardized across countries that is commonly used in GIS studies. | India (admin level = 0), Indian states (admin level = 1), and districts (admin level = 2). State boundaries do not depict the administrative split of the state of Jammu and Kashmir into two UTs – Jammu & Kashmir and Ladakh. District boundaries do not include all districts present in other Indian data sources. 39 and 29 districts present in HMIS and NSS had no matches in GADM, respectively. A detailed list is presented in the corresponding dataset file that can be acquired from the authors. Such districts were excluded from the current analysis. Data were used in all visualizations and for creating unique IDs for cross-referencing districts and states between datasets. |
| URCA (Urban-Rural) | The recent study presented global raster with ordinal | Global raster (.tif) of 1 km<sup>2</sup> resolution and each pixel representing |
| Catchment Areas) raster (38) | catchment area categories based on population densities and nearness to high-density urban centers. Hence, this dataset can help in generating globally standardized rural-urban regions and populations. | catchment area category label. We defined binary categories: rural for CA category label > 7, while urban for CA category label from 1 to 7. Details on category are presented in the ReadMe accompanying the original dataset (38). Data was used in population estimation. |
| Health Management and Information System (HMIS), India (39) | HMIS captures facility-wise health statistics for the entire country. The HMIS Standard Reports publish monthly subdistrict data for several variables for each financial year. | We extracted count data for surgeries (major – those requiring general or spinal anesthesia, minor, major excluding gynecology and ophthalmic operations) c-sections, and institutional deliveries for the period April 2017 – March 2018 and aggregated over subdistricts and months to get district-level annual estimates that were in turn used for state and national aggregations. Variables were partitioned by rural, urban, public and private along. We were interested in rural, urban, and |
|  |  | total values. Data were managed and wrangled on the Google BigQuery cloud platform (40). Some districts that did not match with GADM were excluded from the analysis for consistent calculations and visualization. A detailed list of such districts is presented in the corresponding dataset file that can be requested from the authors. |
| Guilmoto and Dumont (2019) (33) | The article presents state-level c-section proportions relative to institutional deliveries and sampled births using data from National Family Health Survey-4 (2015-16). | Data in the Table 2 of the paper was extracted from the PDF file for comparison with corresponding values from HMIS in the overlapping period. |
| Census-based 2017 population projections (41) | The National Commission on Population reports mid-year state-level population projections for 2011-36. These are arguably the most reliable population projections for India. | We extracted state-level projections from PDF files for 2017 to compare with corresponding raster-based population estimates. Populations for Jammu and Kashmir and Ladakh from the report were combined |
|  |  | under Jammu and Kashmir. |

### 2.2 Data Variables

#### 2.2.1 Rural-Urban population estimation

We used raster-based (rectangular grid of pixels) analysis for creating high-resolution population estimates **(Figure 1)**. The analytical choice was made due to missing district-level 2017 Indian population estimates for rural areas. Hence, district and state-level population aggregations were created for rural areas by partitioning the WorldPop population counts as per the rural-urban dichotomization of catchment areas derived from the multiple catchment area (CA) categories in the URCA dataset (38). We defined CA categories >7 in URCA’s category label classification as rural. Rural populations were estimated as follows – first, the global multi-category URCA raster (1 km^2^ resolution at the equator, pixel = agglomeration category label value) was clipped to the Indian national boundary (admin level-0). Next, the raster was reclassified into two categories: urban (URCA agglomeration and CA labels ≤ 7) and rural (URCA agglomeration and CA labels > 7). Further, the binarized rural-urban CA raster for India was overlayed on the WorldPop 2017 Indian population counts raster (1 km^2^ resolution at equator, pixel = population count) (34). The CA raster was resampled to align the origin and match the extent and resolution of the population raster. The rural population at each pixel was calculated by multiplying the categorical value (1 for rural areas in the rural raster) with the persons per pixel values. Put otherwise, all urban population was weighted by ‘0’ in the rural raster. Similarly, rural CAs were weighted by ‘0’ in the urban raster. Hence, separate rural and urban population rasters were created for India. State (admin level-1) and district (admin level-2) boundaries were imposed on the total, rural, and urban population rasters. Population aggregates (summations) within the boundaries were extracted as district and states population counts. Finally, state-level populations were validated against the Census-based rural, urban, total mid-year population projections for 2017 (41).

**Figure 1:**
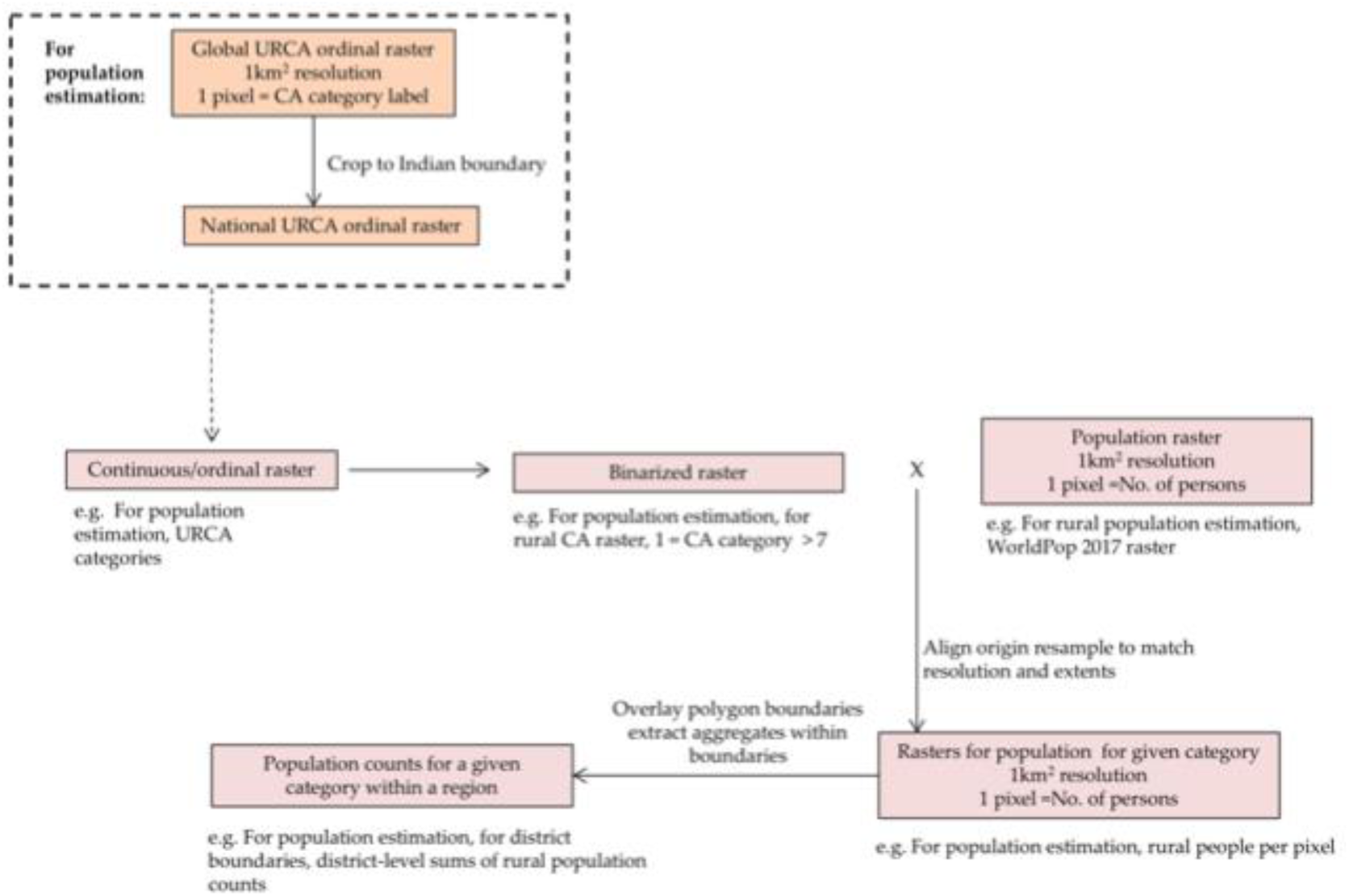
Generic raster-based estimation pipeline used for creating criteria-specific regional population counts.

#### 2.2.2 Surgical and C-section Volumes, Rates/Proportions, and Need

For a given region (district, state, or country-level), the surgical rate was defined as the number of surgical operations (OPs) conducted per 100,000 people in the region. Met need was defined as the ratio of observed surgical rates to the threshold of 5000 surgeries per 100,000 people (9, 42). For the primary analysis, we used the volumes of major surgical OPs (those requiring general or spinal anesthesia) (12) from the HMIS (April 2017- March 2018) for rate and need calculations. For sensitivity analyses, we also calculated the need based on rates for total surgeries (major and minor- not requiring anesthesia) and select major surgeries excluding gynecology (OBGYN) and ophthalmic procedures. For all rate calculations, we used the 2017 raster-estimated populations. C-section proportions w.r.t. institutional deliveries were also calculated. The met need for c-sections was calculated as the ratio of c-sections as the proportion (%) of institutional deliveries relative to the WHO prescribed 10% and 15% thresholds (43). Additionally, we calculated absolute need gaps (i.e. threshold - value) for surgical rates and c-section proportions.

Since this is one of the first instances using HMIS in research, we compared the state-level estimates for two c-section proportions (relative to births and institutional deliveries) from HMIS (January 2015 - November 2016) with corresponding NFHS-4 estimates obtained from (33). This period was chosen to match closely to the data collection period of NFHS-4 (20^th^ January 2015 – 4^th^ December 2016).

### 2.3 Statistical Analysis

All statistical analysis except that for spatial inequality was conducted in RStudio (Version 1.3.1056) using user-created and validated R packages (44). For scrapping values from reports, we used Abbyy FineReader (45), ExtractTable (46), and Tabula (47). Tools used for geocoding are reported above.

#### 2.3.1 Rural-urban comparisons

Given the known skewed data distribution, we used non-parametric pair-wise Wilcoxon tests adjusted for multiple comparisons (Holm-Bonferroni correction) to investigate rural-urban differences for various surgical care variables at state and district-levels. We used the conventional 5% threshold for determining statistical significance. No analysis was conducted for any variables involving select major surgeries due to missing data for urban areas in HMIS.

#### 2.3.2 Validation of estimates

Validation in the form agreement analysis was conducted using Lin’s concordance correlation coefficient (CCC) (48). Agreement was categorized as almost perfect for CCC > 0.99, substantial for CCC ∈ [0.95-0.99], moderate for CCC ∈ [0.90-0.95), and poor for CCC < 0.90 (49). The analysis was conducted for (a) the raster-estimated total, rural, and urban state populations vs. the corresponding Census-based mid-year projections for 2017 (41), and (b) state-level c-section percentages relative to births and institutional deliveries from HMIS (January 2015-November 2016) vs. corresponding NSFH-4 based values (33).

## 3. Results

### 3.1 Raster-based population estimates

The district and state-level 2017 population aggregates for rural, urban, and total populations can be requested from the authors. The state-level total and rural estimates showed almost perfect agreement with corresponding RGI Census projections, while urban estimates had a substantial agreement as revealed by Lin’s concordance correlation coefficient **(Appendix B)**.

### 3.2 Surgical volumes, rates, and need

At the national level, rates (per 100,000 people) of the total, major (requiring general or spinal anesthesia), and select major (excluding gynecology, ophthalmic, and other procedures) surgical operations (OPs) in rural regions were 1274, 341, and 100, respectively. Significant differences between rural and urban of varying sizes were found at state and district-level comparisons for total and major rates, while no comparisons were conducted for select major OP rates due to limited data for urban regions **(Appendices C & D)**. We found higher surgical rates in rural regions compared to urban regions at the state level. However, this might be an artifact due to the limited upscale of HMIS in urban regions or misclassification of certain regions (see **Discussion Section 4.3**).

For the primary analysis, the met surgical need was defined as the ratio of the rate of major surgical OPs to the threshold of 5000 surgical procedures. Nationally, rural regions’ met surgical need was at 0.0681 or 6.81%. For rural regions, most districts and states fell under the 0.20 (or 20%) mark for the met need w.r.t major surgeries **(Figures 2A-B)**. In certain states such as Arunachal Pradesh, Rajasthan, and Maharashtra, a small number of districts with a higher met need ratio skewed the met surgical need at the state level in an upward direction although a greater number of districts had low surgical rates. Further, some differences between inference guided by state and district resolution maps arise due to skipping the HMIS districts that were not matched with GADM in the district-level map. However, these districts do contribute to the state-level estimates thereby altering the level of met need in the state-level map, e.g. Telangana and Sikkim.

**Figure 2:**
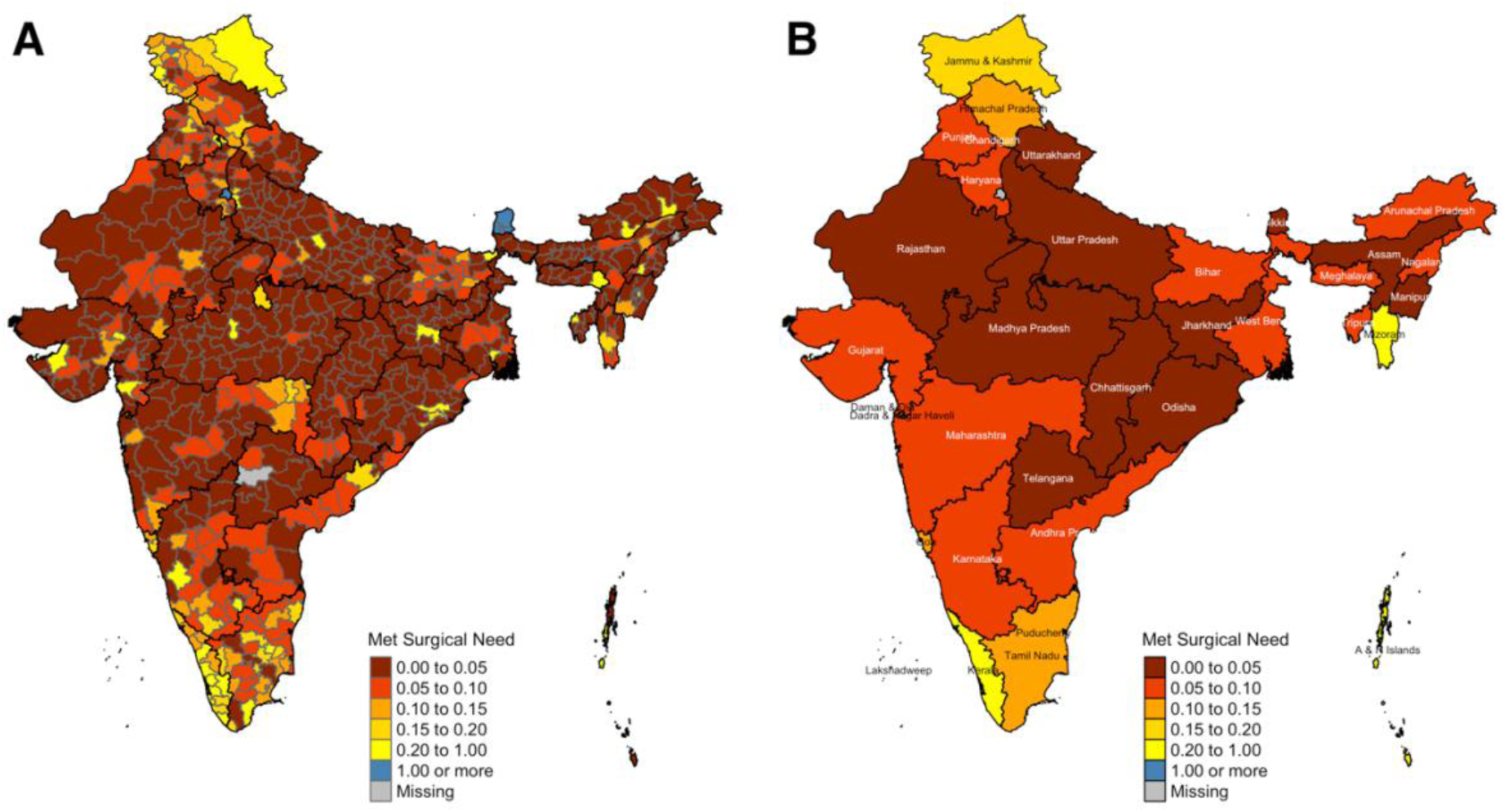
Geographic variations in rural met need for major surgeries at – A) district and B) state-levels.

In district **(Figure 3A)** and state-level **(Figure 3B)** comparisons, surgical met need for major OPs differed significantly between rural and urban regions with small-to-moderate effect sizes **(Appendices C & D)**.

**Figure 3:**
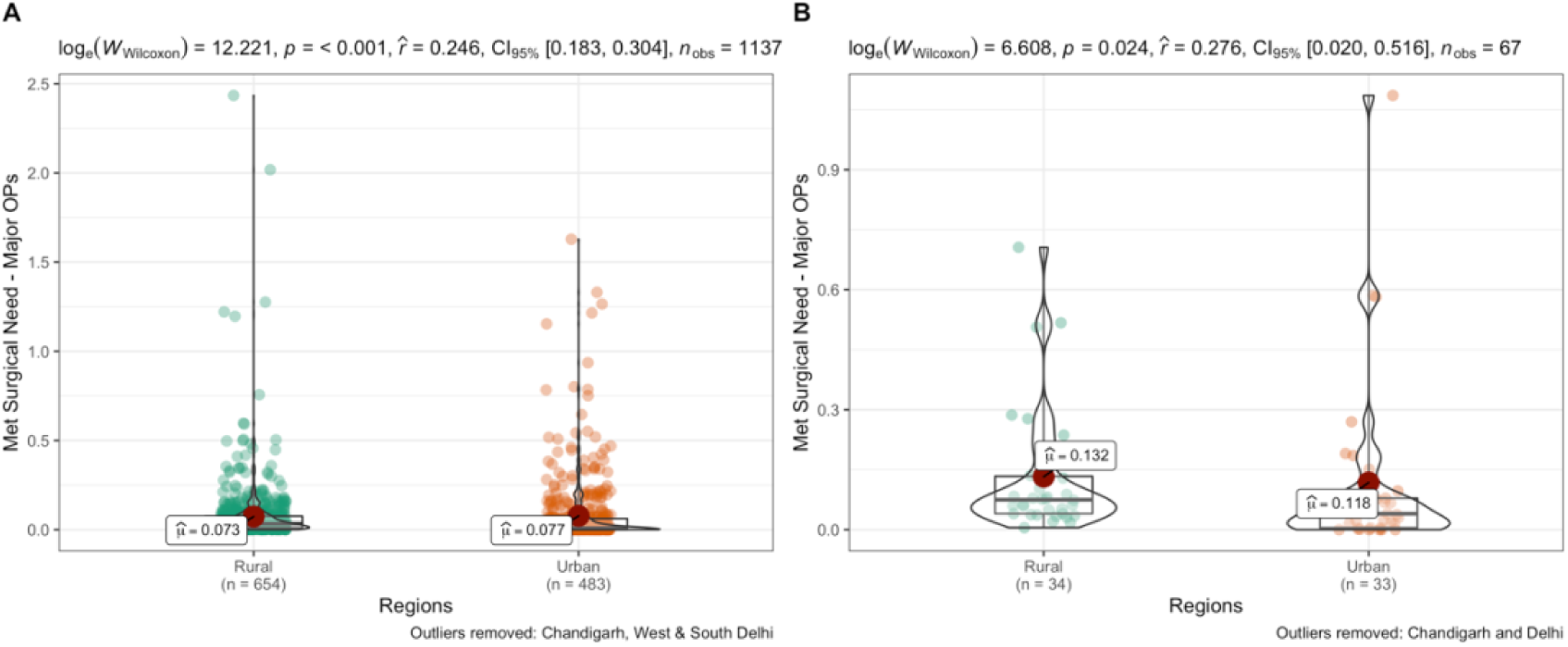
Rural-urban differences in met need for major surgeries at A) district and B) state-levels.

The rural-urban differences held up for other variables such as met need w.r.t total surgical OPs, absolute surgical need gaps w.r.t total, and major OPs **(Appendices C & D)**. As mentioned before, the rural-urban comparisons should be considered with caution.

### 3.3 C-section proportions and need

Nationally, 13.57% of all rural institutional deliveries were c-sections falling within the WHO prescribed 10-15% range. Met c-section need w.r.t 10% threshold was at 1.36. For rural regions, almost all districts and states in southern India depict satisfactory performance (met c-section need >1) pointing to a north-south divide with Bihar, Uttar Pradesh, and Rajasthan requiring attention **(Figures 4A-B)**.

**Figure 4:**
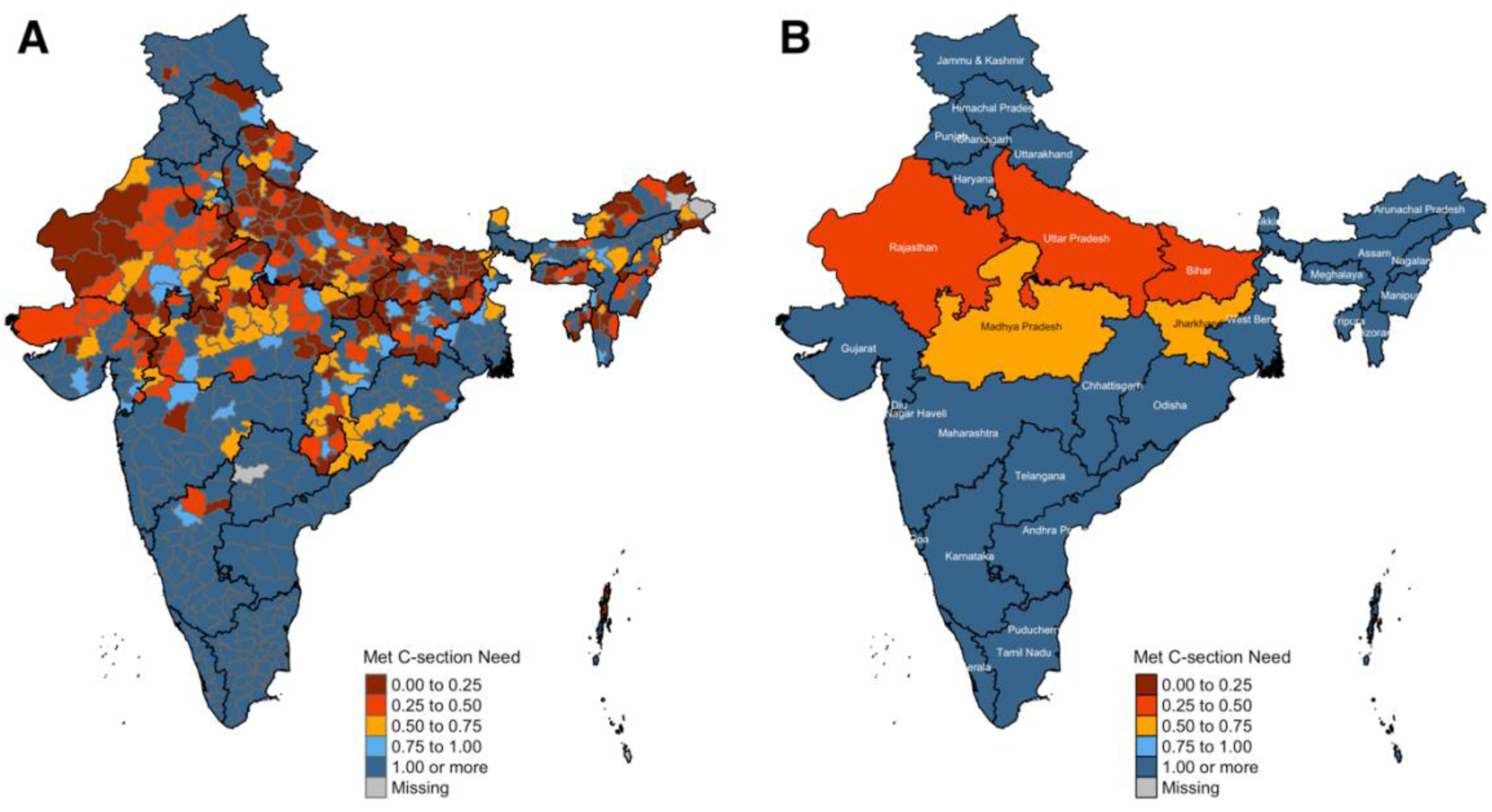
Geographic variations in met c-section need (at 10% threshold) for rural regions at – A) district and B) state-levels.

At the national level, 33.92% of institutional deliveries were c-sections in urban areas depicting an excess. State and district-level comparisons between rural and urban regions had significant small-to-moderate sized differences for the met need of c-sections at 10% threshold **(Figures 5A-B)**, proportion out of institutional deliveries, met need as per 15% threshold, and absolute gaps (excess or deficit) as per both thresholds **(Appendices C & D)** with high values for urban regions.

**Figure 5:**
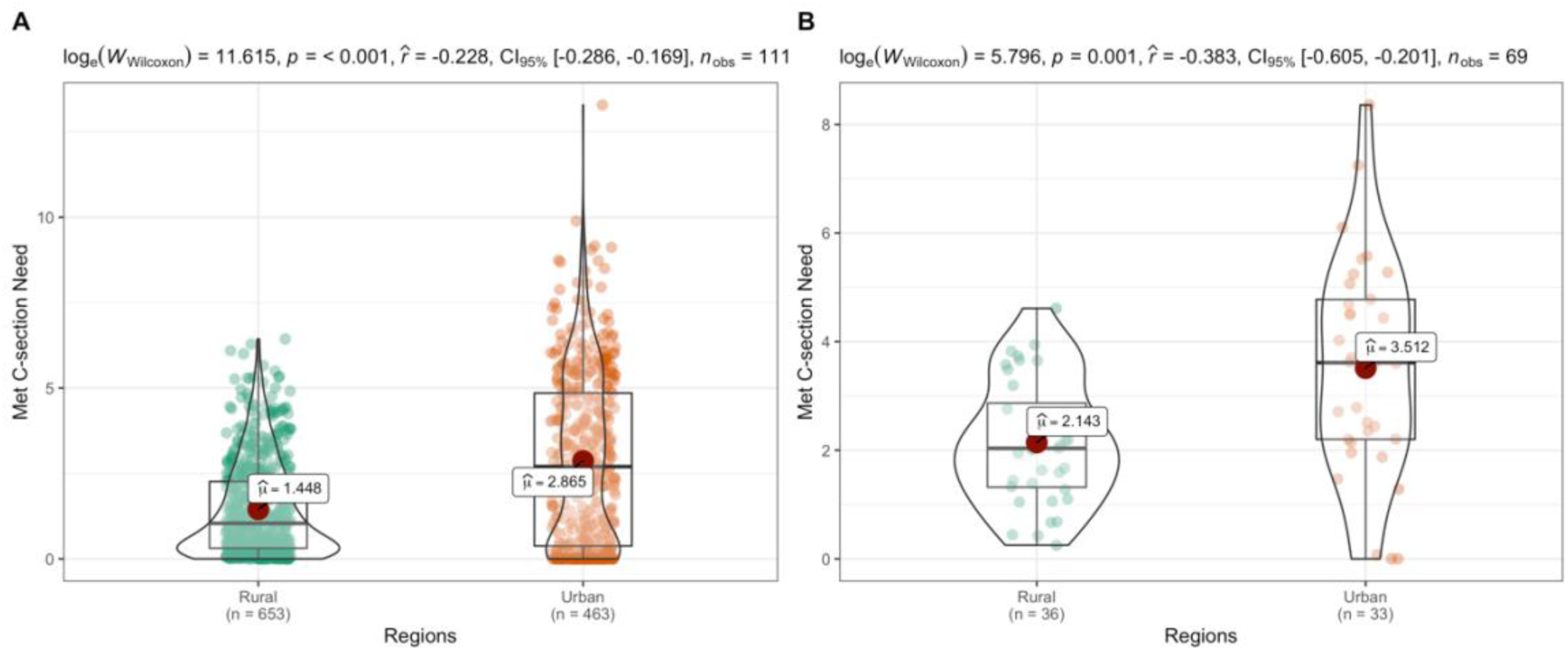
Rural-urban differences in met c-section need (at 10% threshold) at A) district and B) state-levels.

### 3.4 Validation of c-sections from HMIS vs. NFHS

For the overlapping period of January 2015 to November 2016, the state-level population and institutional c-section proportions showed poor agreement between HMIS and NFHS-4 **(Appendix E)**. Andhra Pradesh and Telangana that were formed in 2014 following an administrative split in the parent state of Andhra Pradesh were identified as outliers. The agreement improved after the removal of outliers with population c-section percentage reaching a moderate level.

## 4. Discussion

The current study primarily focused on rural India for the year 2017-18. We synthesized volumes, rates or proportions, and need for surgeries and c-sections for over 660 districts in 36 states and union territories for the rural populations. Briefly, we found that the surgical rates in rural India fell short of the met need benchmark for major surgeries. While the findings for c-sections are varied and complex. The validation steps throughout the primary analysis and the adjunct sensitivity analysis confirm the utility of our methodological approach and the robustness of our findings.

### 4.1 Contextualizing the study findings

Our national-level rural surgical rate estimates fall within the range of LMIC values presented under Indicator 3 of the Global Indicators Initiative (12). The total and major surgical rates were higher than the LCoGS associated modeled estimates for India (10). However, this model is also known to underestimate surgical rates in other South Asian countries such as Myanmar and Sri Lanka (12). The rural rates of select major surgeries (excluding OBGYN, ophthalmic, and other procedures) for Andhra Pradesh (170 per 100,000 people) and Telangana (106 per 100,000) were quite under the previously known 259 per 100,000 beneficiaries from an insurance claims study in the region (11). This difference could be attributed to populations covered (beneficiaries at pre-dominantly private hospitals for (11)), surgical OPs considered, and study periods (data from mid 2008-12 for (11)). Our national estimates for total and major surgical rates in rural regions are less than the recently projected national population estimate of 3,646/100,000 (17). The projection in (17) was created based on the electronic medical records of surgical uptake in a well-characterized urban UHC cohort in Mumbai, Maharashtra. Our HMIS-based estimates (total: 1,851/100,000 major: 1,003/100,000) for the district of Bruhan Mumbai (urban) fall short of the UHC cohort estimate (4,642/100,000). Investigating the reasons underlying these differences would require assessment of data coverage and completeness for HMIS and further breakdown by surgical conditions to better match the inclusion of the surgical condition across studies. Our population-based met need calculations cannot be compared with small-scale household surveys investigating lifetime surgical uptake (50) or studies using self-report instruments such as SOSAS (18, 19).

The c-section proportion estimates match with other similar nationwide studies using representative household surveys (see **Appendix A**). The north-south divide in the met c-section needs observed in our analysis matches with that presented by Guilmoto and Dumont (33). The met need for c-sections is much higher than that for overall major surgeries. This could be due to the following reasons: a) c-section scale-up in LMICs has been encouraged under the OBGYN, maternal and childcare programs (51) due to known deficits in the early 2000s (52) and b) India has observed an increasing incidence of unnecessary c-sections with large proportions of such procedures conducted in private facilities (see **Appendix A**). Regardless of the underlying reasons, it is important to note that the use of c-sections as a proxy for met need can be problematic.

### 4.2 Implications for policy and practice

Two major policy and practice offshoots from the LCoGS research include the proposals for the global surgical care indicators (13) and NSOAPs (8). The current study contributes to both these proposals by providing nationwide estimates for Indicator 3 that can be used for baseline situational analysis for NSOAP. Further, our methodological workflow from data assimilation to estimation and visualization including the use of adaptable inexpensive tools can act as a blueprint for situational analysis essential for NSOAP development. Considering that several LMICs currently lack NSOAPs, the blueprint has high translational value for researcher-policymaker partnerships globally. In India, the first-ever nationwide findings for rural populations can garner interest from the national and state government stakeholders. The government buy-in can encourage policymakers to put the current research findings to use. Engagement with policymakers using current analysis from eclectic sources could create avenues for data sharing from HMIS and PMJAY insurance claims databases for researchers.

### 4.3 Study strengths and limitations

To our best knowledge, this is the first study investigating nationwide surgical need in India. More importantly, we generated the estimates for the rural populations that need them the most. The homogenized dataset of district-level surgical care variables has high value for researchers and policymakers alike. Our analytical pipelines are open-source and minimally expensive. Hence, they can be easily employed by other students and researchers working under financial resources constraints. Specifically, our data assimilation pipeline can be easily translated to other countries to generate surgical care variables datasets at subnational resolution. The raster-based population estimation pipeline can be extended to any global region to get general and sex-age specific population counts disaggregated by rural-urban partitions for a given administrative or geographic unit without requiring GIS expertise or high computing power. The current study marks one of the first instances using HMIS data for research and we provide a comparison of c-section variables against NFHS. Typically, subnational studies for India focus on states. However, within-state variations identified in this study demonstrate the need and usefulness of high-resolution estimates.

We acknowledge that the study has several limitations. First, like any secondary data analysis, our estimates inherit the known limitations of the parent datasets. For instance, any known limitations of the global URCA raster (38) apply to population estimation analysis. HMIS analytical reports have shown greater estimates for generic health system variables such as immunizations, institutional deliveries, child and maternal mortality rates, etc. when compared to sample surveys such as Annual Health Survey and District-Level Household Survey, etc. (53) In this study, we compared the c-section proportions against NFHS-4 for a similar time window. While the agreement was classified to be ‘poor’, the value of Lin’s concordance correlation was quite large, particularly after removing the outliers. The limited agreement could be due to exclusion of several districts in Telangana that did not match with GADM, inexact matching of the periods (i.e. 20^th^ January 2015 to 4^th^ December 2016 in case of NFHS-4 (54) was matched with January 2015 to November 2016 from HMIS due to presence of only monthly data), differences in the definition of births in case of population proportion (presumably all births were considered for NFHS-4 (33) while only live births were considered for HMIS), etc. Even so, data quality as measured by completeness, actual vs. reported coverage, etc. for other non-surgical variables has been studied for HMIS (55, 56). Regardless, we promote the use of HMIS data as it is relevant for local health planning in LMICs and increased use in academic research can provide feedback on potential data issues. State or district-wise differences in underlying datasets can further impact our subnational estimates. Second, our analysis of surgical volumes, rates, or need did not consider the classification of surgical operations by underlying conditions or patient demographics since we used the district-level aggregates from HMIS. Further analysis would be possible with more extensive and transparent data sharing from HMIS. Third, the current HMIS dataset was inflated with NA (not available) and zero values. We included the zero values to avoid investigators’ bias. However, these data issues can make estimates unreliable. Hence, the observed low values could be erroneous. In the future, data imputation techniques can be used to model surgical volumes to correct bias due to excess zeros.

### 4.4 Implications for future research

The current study could inspire research along multiple lines. For India, modeled estimates generated through meta-analyses, the release of new data from HMIS, or using other data sources such as the PMJAY insurance claims database could help. Creating dis-aggregated estimates for public vs. private surgical facilities, adult and pediatric populations, socio-economic status of surgical care seekers and facilities in rural vs. urban regions is critical for comprehensive understanding. The resolution of can be enhanced to the sub-regional surgical facility-level if relevant data are available by HMIS. Formal distributional and geospatial inequality analyses should be conducted for various surgical care variables to better understand the distributional and spatial patterns for directing policy interventions. Uncertainty analysis (57) accompanying small area estimation or population microsimulations (see (58) for methodological review) can enhance the robustness of estimates. The current estimates should be validated by facility-level randomized assessments in select Indian districts collecting data on surgical care access dimensions similar to those conducted in Ghana (59) and Uganda (60). Beyond India, the methods and tools proposed in the current study have high translational value for global surgery research. Our methodological approach relies on HMIS data that is common across LMICs and hence can be easily adopted in similar settings such as the neighboring South-Asian countries or those beyond. We also plan to develop an application interface for our analytical pipelines to improve access to research tools.

## 5. Conclusion

We present the first-ever high-resolution nationwide surgical volumes, rates, and need estimates that can inspire the initiation of NSOAP development for India. We developed methods that have high translational value for synthesizing similar surgical care access estimates at subnational resolution in other low-and-middle-income countries. The Indian estimates can be further improved by overcoming the limitations of our study and with the availability of more data. Future studies should extend our findings to include other surgical care indicators.

## Data Availability

All data produced in the present study are available upon reasonable request to the authors.

## Acknowledgments

This monograph was adapted from the thesis submitted by Siddhesh Zadey in partial fulfillment of the requirements for the degree of Master of Science in the Duke Global Health Institute in the Graduate School of Duke University. We thank Dr. Swati Sonal, Dr. Sweta Dubey, and Mr. Pushkar Nimkar from the <u>Association for Socially Applicable</u> <u>Research (ASAR), India</u> for their contributions to the findings presented in the monograph. We also thank Dr. Catherine Staton and Dr. Tamara Fitzgerald from Department of Surgery, Duke School of Medicine and Duke Global Health Institute (DGHI) for their insightful comments, feedback, and counsel. We thank Drs. Hampus Holmer, Gnanaraj Jesudian, Rani Bang, and Nakul Raykar for helpful discussions. We acknowledge support from everyone at ASAR, DGHI and GEMINI Research Lab for their support.

## Declaration

Ethics approval and consent to participate: Not applicable Consent for publication: Not applicable

Availability of data and materials: The dataset can be requested from the authors. Competing interests: The authors declare no competing interests.

Funding: The study did not have any primary research funder or sponsor. From 2019-2021, SZ was supported by Duke Global Health Institute Merit Scholarship and Assistantship; JV is supported by multiple grants at DGHI and Duke University School of Medicine.

## Appendix A

Summary of studies on c-section (CS) proportions in recent years based on nationally representative surveys

| Authors | Study description | Estimate |
| --- | --- | --- |
| Ologunde et al. (2014) (61) | Cross-sectional analysis of facility-based data on LMICs under WHO Situational Analysis Tool to Assess Emergency and Essential Surgical Care, including 171 Indian facilities, of which 110 provided data on CS deliveries. | % CS births of total births in 2008- 8.1% |
| Neuman et al. (2014) (62) | Cross-sectional analysis of data collected during cluster-randomized controlled trials (c-RCTs) in Asian countries, including India (rural - Jharkhand and Orissa, 2005-08; urban - Mumbai slums, 2006-09) assessing CS prevalence and determinants in private and public facilities. | Cluster-level estimates of % CS births of total live births for 2011-Rural-Public: 15%<br>Rural-Private/Charitable: 5%<br>Urban-Public: 15%<br>Urban-Private/Charitable: 18% |
| Singh et al. (2018) (63) | Secondary analysis of DLHS-4 for comparing CS births across public and | National-level estimates of % CS births of facility-based |
|  | private facilities. Authors present analysis of trends and sociodemographic and health service utilization correlates but do not depict inter-state or district differences. | deliveries-Private facilities: 13.7%<br>Public facilities: 37.9%<br>Urban areas: 28.6%<br>Rural areas: 19.5% |
| Guilmoto et al. (2019) (33) | Secondary analysis of NFHS-4 for CS births w.r.t institutional deliveries and population-level births. The authors present an analysis of trends, state and district differences, and sociodemographic correlates. | National-level estimates of % CS births of-Population sampled births: 17.2%<br>Institutional deliveries: 21.8%<br>District and state-level estimates are given. |
| Bhatia et al. (2020) (64) | Secondary analysis of NFHS-3 and -4 for CS births in public and private facilities.<br>Authors present analysis of trends, state-level differences, and sociodemographic and health service utilization correlates but do not depict district-level differences. | National-level estimates of % CS births of institutional births-NFHS-3 (2005-06)<br>Public facilities: 15.2%<br>Private facilities: 27.9%<br>NFHS-4 (2015-16) |
|  |  | Public facilities: 11.9%<br>Private facilities: 40.9%<br>State-level estimates are presented. |

## Appendix B

Agreement analysis to validate 2017 state-level estimates for A) total, B) rural, and C) urban populations.

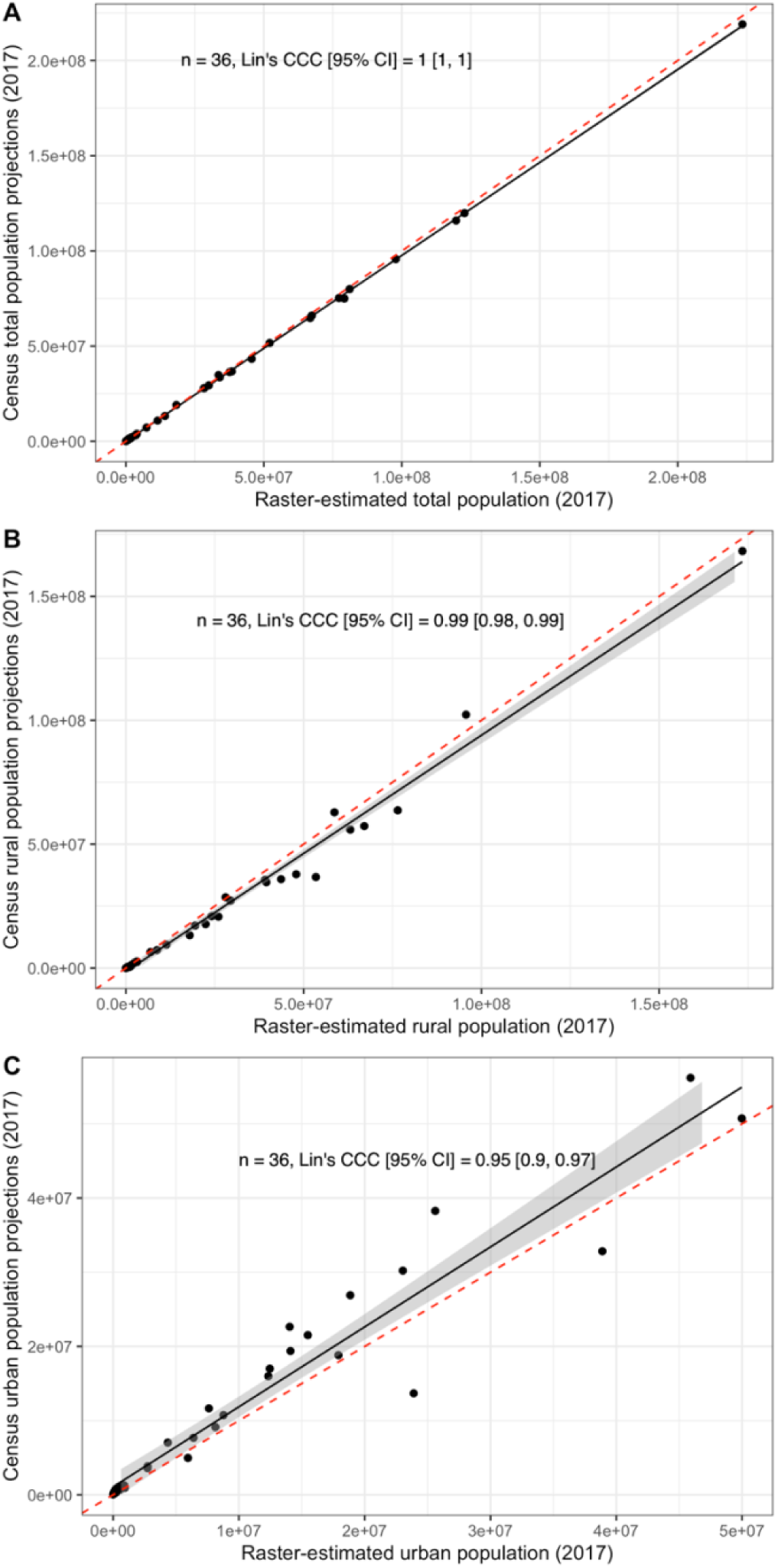

## Appendix C

State-level rural-urban differences for surgical care variables.

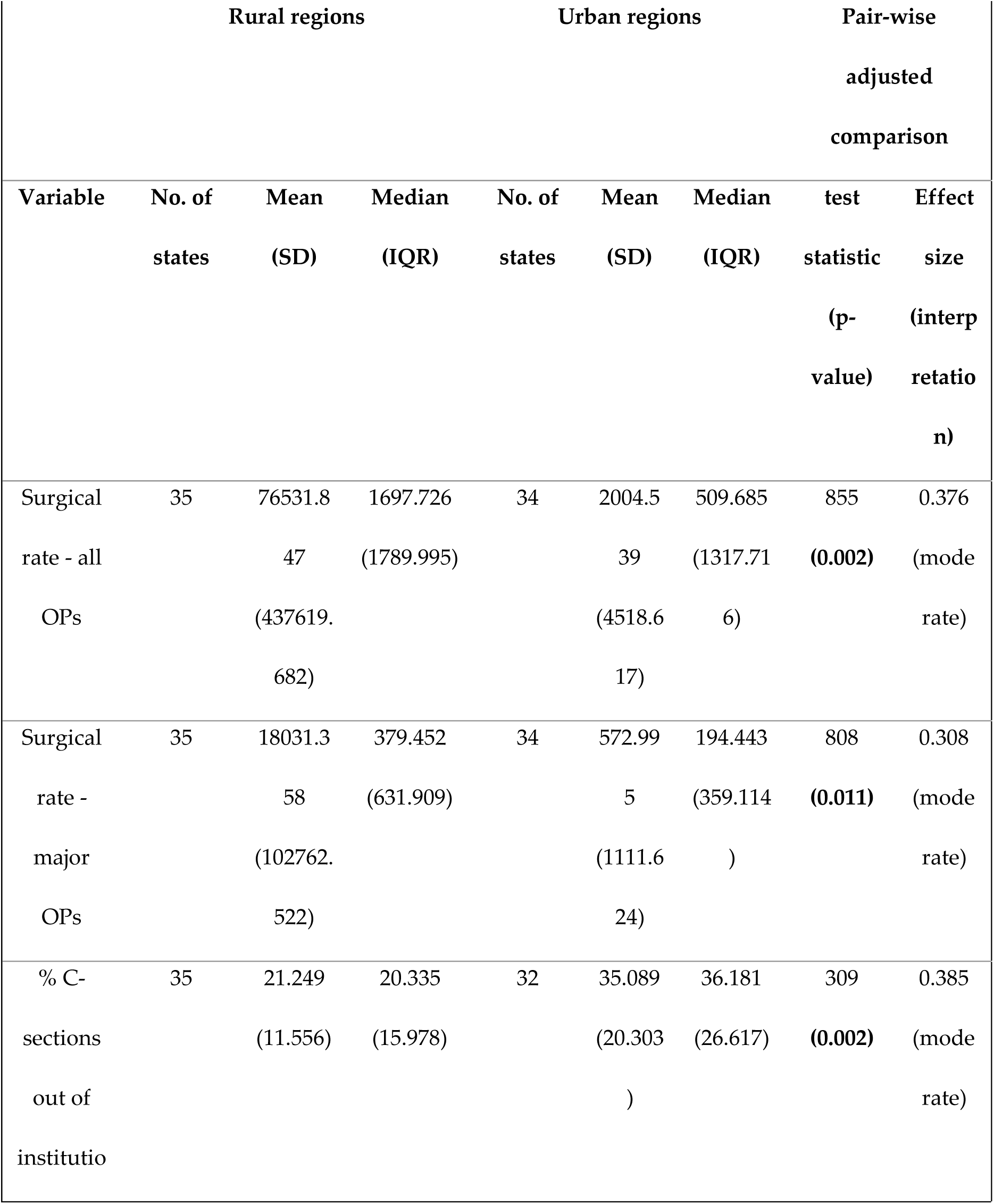
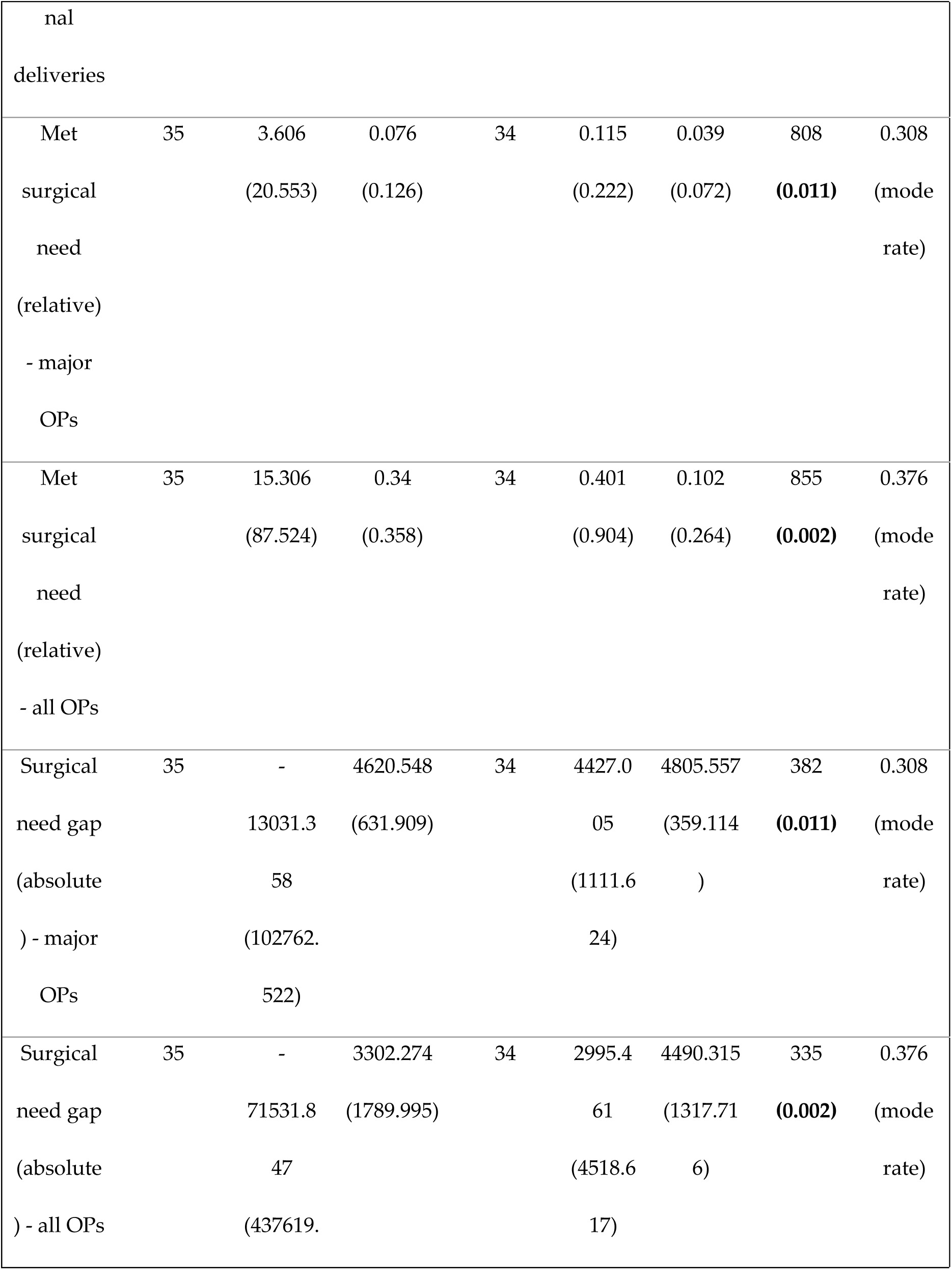
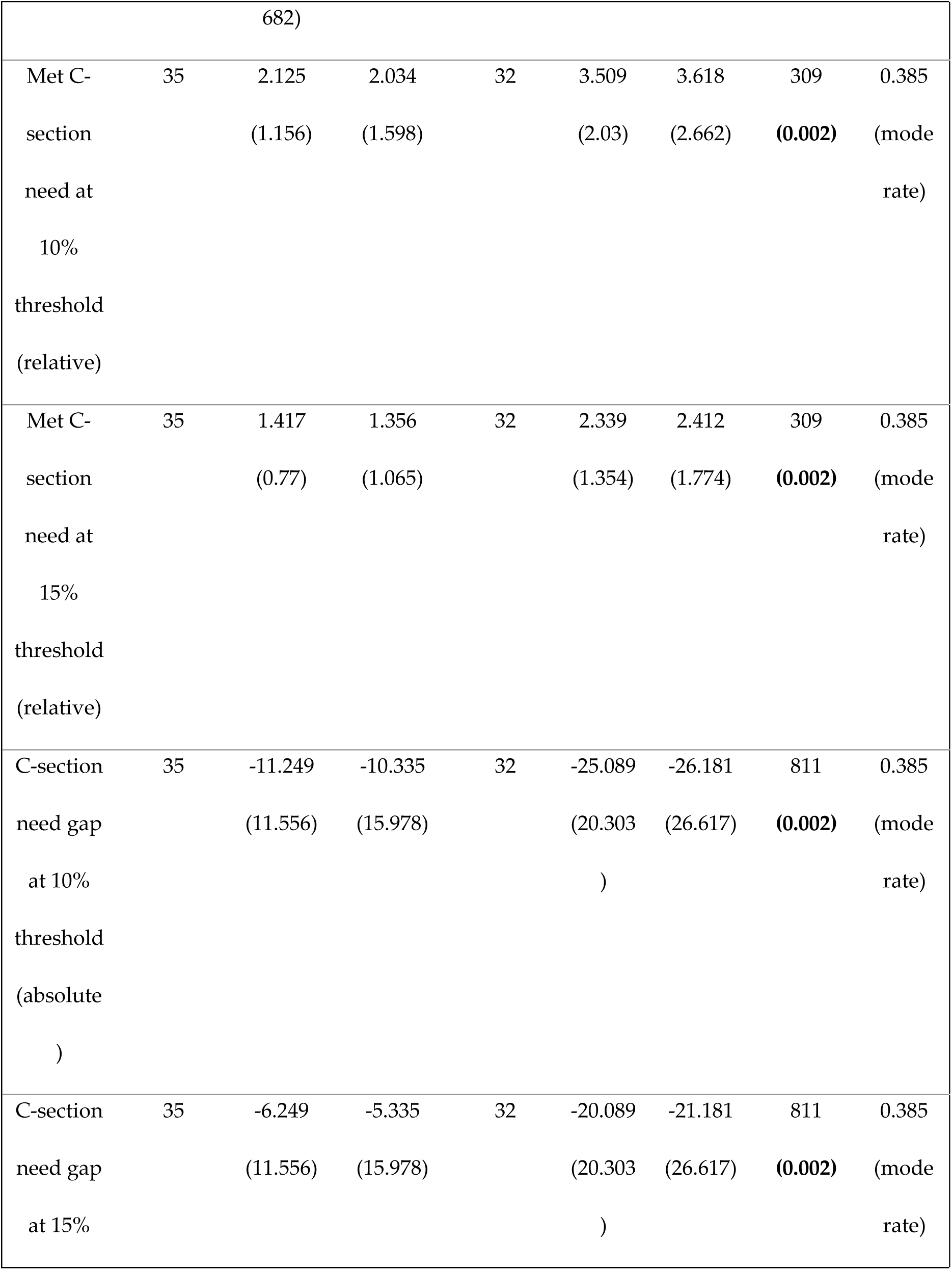
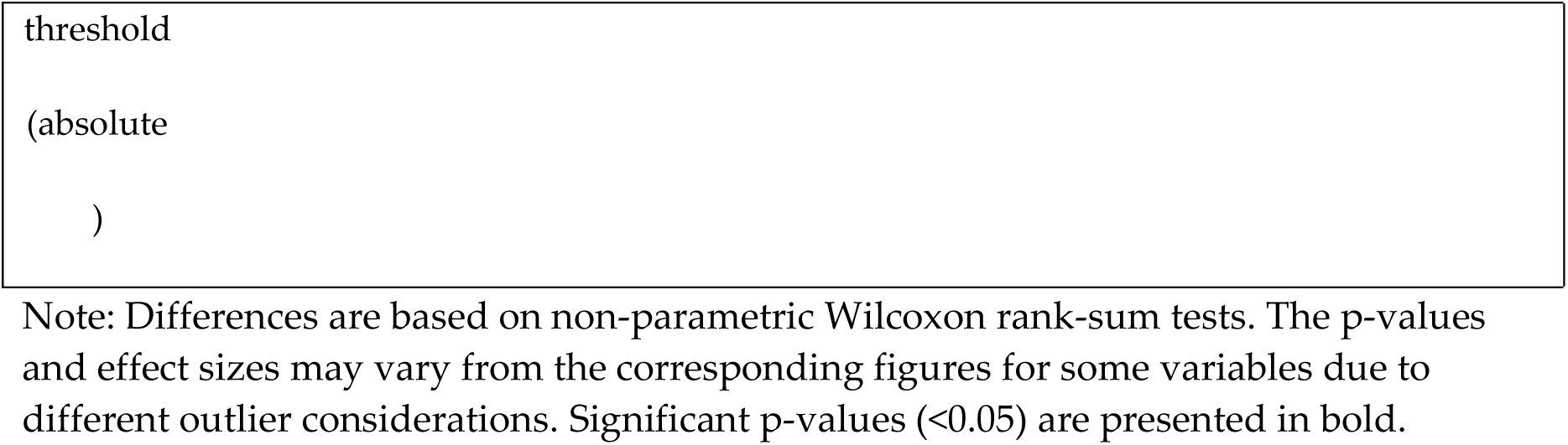

## Appendix D

District-level rural-urban differences for surgical care variables.

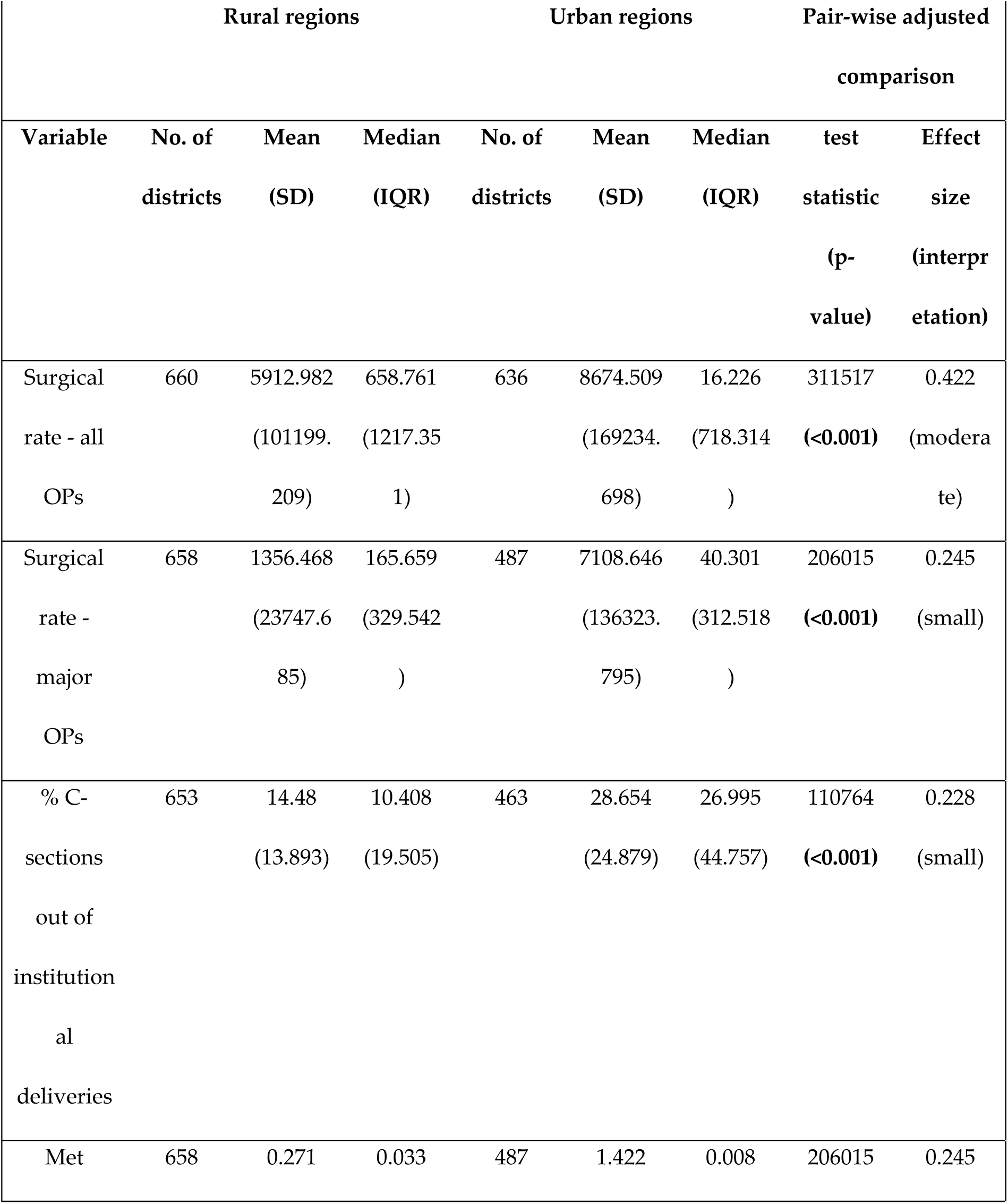
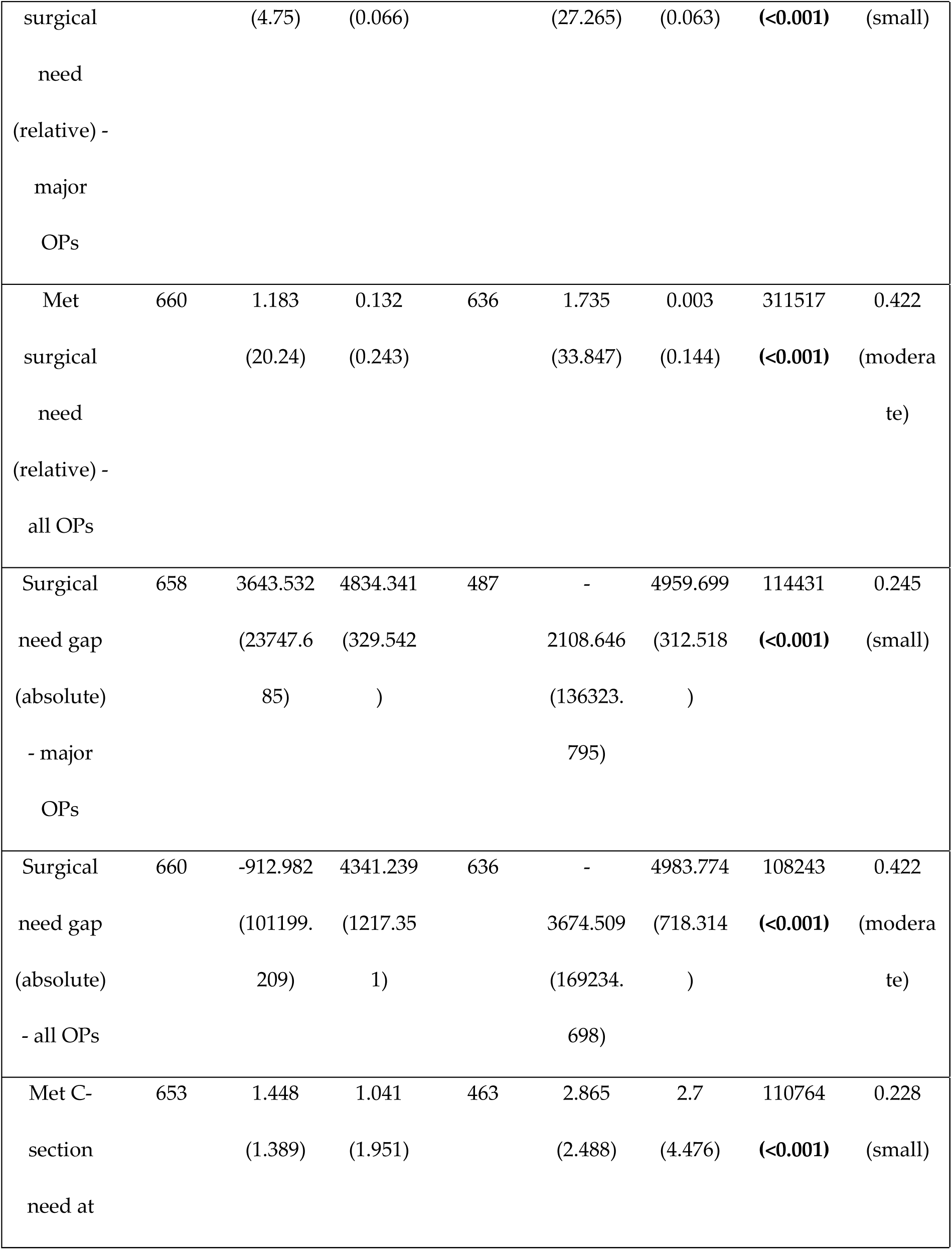
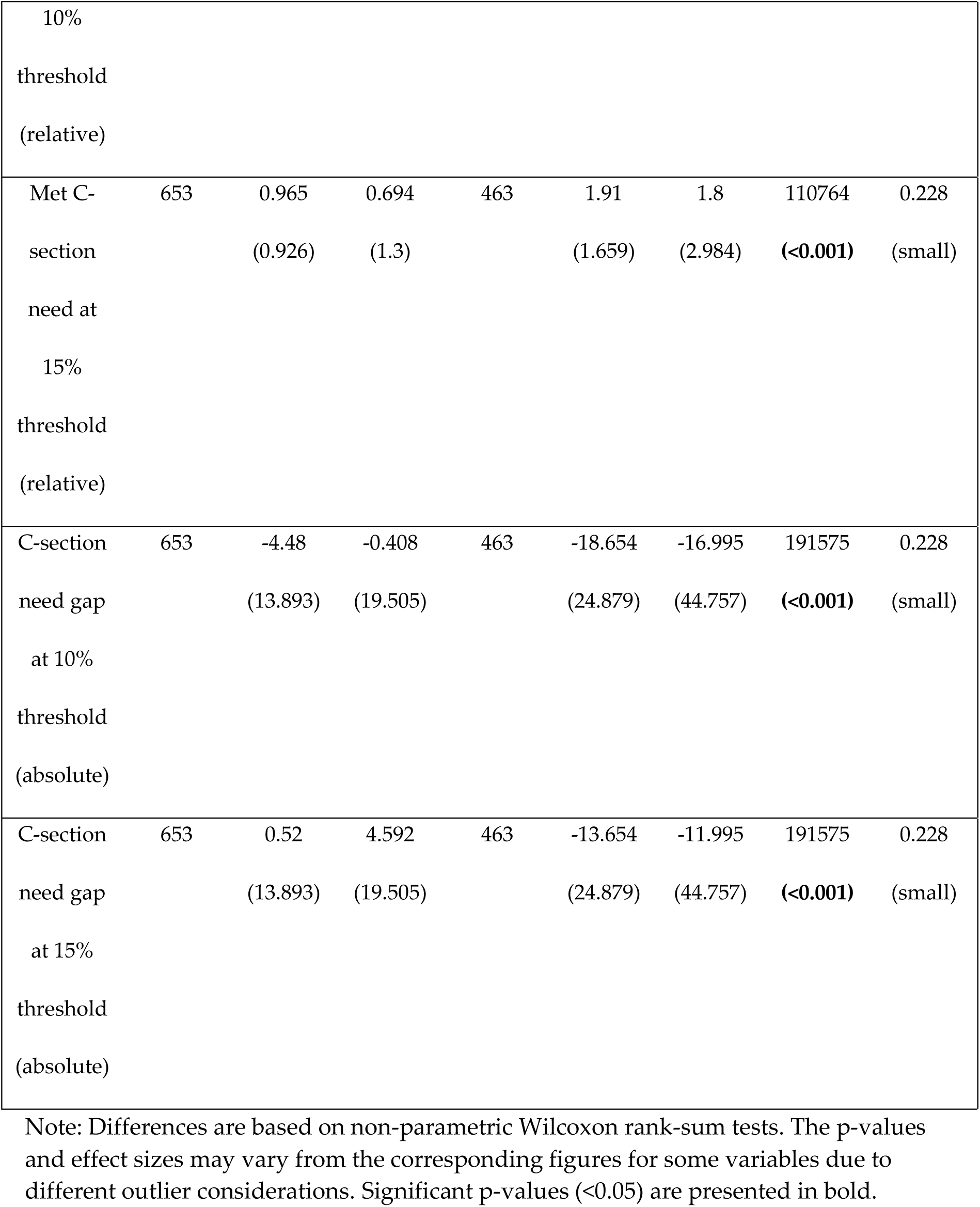

## Appendix E

Agreement analysis to validate state-level HMIS values against corresponding NFHS-4 based estimates for c-sections as A) % institutional deliveries, B) % births.

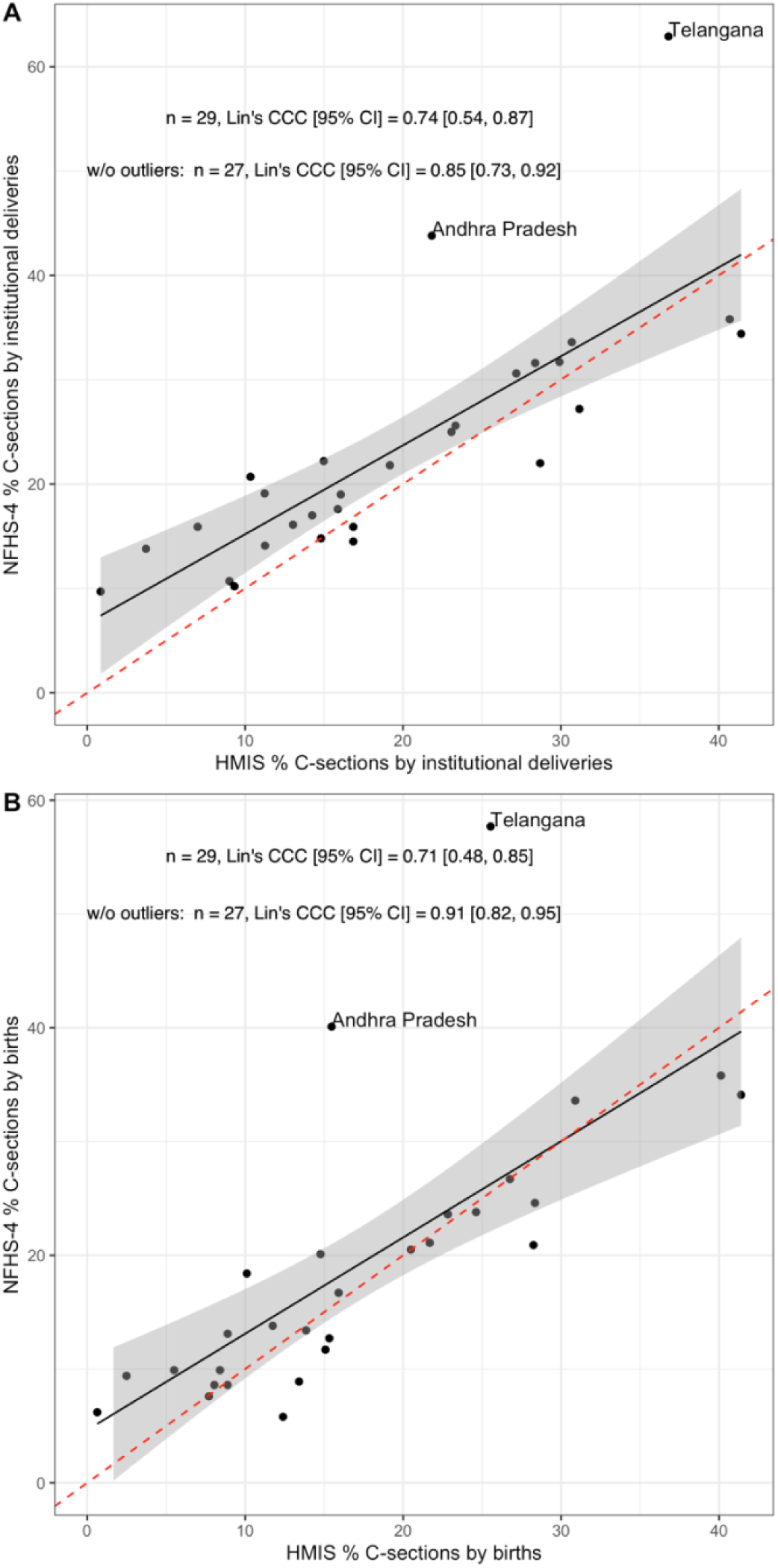

